# Kinesiotaping, single-shot chest wall regional anaesthesia, and Incentive Spirometry for Rib Fracture Management in the Emergency Department: A Systematic Review and Meta-Analysis

**DOI:** 10.64898/2026.08.17.26360233

**Authors:** Sophie Lenihan, Marc Barr, Kate Coates, Louise Kedroff, Ceri Battle, Vittoria Sorice, Mark A Faghy, John Edwards, Diana Papaioannou, Tracey Young, Ines Rombach, Edward Carlton, Steve Goodacre, Nick Mani

## Abstract

**Background:** Pain management post-rib fractures is often difficult. High pain levels can lead to altered respiratory mechanics and delayed complications such as poor mobility. Whilst as-needed Opioids are the mainstay of treatment, the potential negative side effects have led to research into alternatives such as kinesiotaping, single-shot chest wall regional anaesthesia, and incentive spirometry.

**Methods:** A systematic review was undertaken using Medline (Ovid), Emcare, CINAHL, and the Cochrane Library. Article review and selection were undertaken by two independent reviewers using Covidence. Quality was assessed through the Mixed Methods Appraisal Tool (MMAT). Where appropriate, meta-analysis was undertaken using R studio with a REML random effects model. Forest plots were completed, and Higgins’ I^2^ and Chi^2^ were calculated.

**Results:** Kinesiotaping demonstrates a reduced pain score than medication alone (SMD: -1.87, 95% CI [-2.65, -1.08]), as did single-shot chest wall regional anaesthesia (SMD: -0.79 [-1.15, -0.01]). The regional anaesthesia group had lower opioid consumption (SMD -0.84 [-2.18, 0.50]) and reduced length of hospital stay (SMD: -0.18 [-0.39, 0.03]) but no change in the risk of complications (RR: 0.92 [0.36, 2.36]). The incentive spirometry group had an increased risk of complications (RR: 3.35 [0.68, 16.44]); however, the causative effect could not be inferred due to significant confounding variables.

**Conclusions:** Low-to-moderate certainty evidence suggests that kinesiotaping and single-shot chest wall regional anaesthesia may reduce pain in adult emergency department patients with rib fractures. However, evidence is insufficient to show a clear benefit for opioid reduction, length of stay, or complications. The current evidence does not support routine use of incentive spirometry in this setting, but the evidence is severely confounded by baseline injury severity in the current published studies.

**Key Messages:** *What is already known on this subject:*

□ Rib fractures are a common presentation to the Emergency Department and are associated with significant pain and respiratory complications.
□ Opioids are the mainstay of treatment but carry significant side effects, driving the need for multi-modal, opioid-sparing analgesia strategies.

*Research Question:*

- Are kinesiotaping, incentive spirometry and single-shot chest wall regional anaesthesia effective adjuncts to standard analgesia in adult emergency department patients presenting with rib fractures following blunt chest wall trauma?

*What this study adds:*

□ Kinesiotaping and regional single-shot chest wall regional anaesthesia including serratus anterior plane block and erector spinae plane block, may reduce pain compared to medication alone.
□ single-shot chest wall regional anaesthesia is associated with reduced total opioid consumption and reduced length of hospital stay in this population.
□ Current evidence for incentive spirometry is confounded by baseline injury severity, so routine use in the emergency department cannot be supported on the basis of high-quality evidence.

## Introduction

Rib fractures are involved in 15% of all trauma cases within UK emergency departments (ED) [1]. They represent a substantial economic burden to the National Health Service. Although there are no current national costings published, mean hospital costs per patient frequently exceed £25,000, primarily due to associated complications such as pneumonia and the need for critical care [3].

While often dismissed in younger patients, severe pain from rib fractures impairs mobility, cough efficacy, and respiratory mechanics, predisposing patients to atelectasis, hypoxaemia and pneumonia, which develops in approximately 10% of cases, often within 2 weeks post-injury [1]. Mortality risk escalates significantly in older adults, with an odds ratio of 2.5 for in-hospital death (95% CI 2.3-2.8) in those over 64 years when severe rib fractures are present, independent of other trauma [4].

Achieving effective analgesia remains elusive, contributing to adverse outcomes [5]. Notably, up to 18.6% of UK rib fracture patients experience unplanned ED re-attendance due to inadequate pain relief and 62% and 57% of patients reporting chronic pain and disability at 3 months post-injury, respectively [1]. Despite recommendations for a multi-modal approach, opioid-based management remains a mainstay of acute treatment, this is concerning, given the contribution of ED prescribing to opioid dependence [8] and the known side effects of opioids, such as respiratory depression, falls, and delirium, which may contribute to post-injury complications [8].

This has led to research into alternative and adjunctive interventions. Regional analgesia, such as the Erector Spinae Plane Block (ESPB) and Serratus Anterior Plane Block (SAPB), has shown promise in reducing pain and opioid consumption after cardiothoracic surgery [5, 10]. Kinesiotaping, though its mechanism is unclear, is thought to exert benefits through a combination of skin-lifting effects that may improve microcirculation, modulation of nociceptive input, enhanced fascial and muscular support, and improved thoracic mobility mechanisms. Collectively, these may reduce pain and support respiratory function in patients with rib fractures to reduce post-injury [11, 12]. Finally, incentive spirometry is proposed to reduce pulmonary complications by encouraging full lung expansion [13].

Despite individual promise, no systematic synthesis evaluates these interventions, kinesiotaping, incentive spirometry and single-shot chest wall regional anasthesia (ESPB/SAPB), against opioids for pain control, complications, opioid sparing, mortality, and hospital stay specifically in ED-presenting adults with rib fractures. This systematic review and meta-analysis address this gap to inform evidence-based ED practice.

## Methods

### Search Strategy and Selection

This systematic review adhered to the Preferred Reporting Items for Systematic Reviews and Meta-Analysis (PRISMA 2020) guidelines [15] and was registered with PROSPERO (CRD42024586150).

Medline (Ovid), Emcare, CINAHL, and the Cochrane Library, alongside a grey literature search using Google Scholar, were searched on 12th December 2024; an updated search was also completed on 8th June 2026. Search terms included combinations of ((“Rib Fractures” OR (rib* AND fracture*) OR “Chest wall injury*”)) AND ((“Emergency Department*” OR “Accident and Emergency”)). The full search strategy is available in **Supplementary File 1**.

Covidence software was used for screening. Two independent reviewers screened all titles and abstracts (SL and MB), followed by full-text reviews. Disagreements were resolved through mutual consensus.

Due to a lack of high-quality RCTs, studies were included if they were observational studies (prospective or retrospective), RCTs, quasi-RCTs, or cross-sectional studies with at least 10 participants. Eligible studies involved adult patients (>18 years) with clinically or radiographically confirmed rib fractures from blunt trauma. The interventions of interest were kinesiotaping, single-shot chest wall regional anaesthesia, or incentive spirometry, compared to usual care (predominantly opioid medication alone). We excluded systematic reviews, non-English papers, and studies involving patients with significant polytrauma (e.g., abdominal injuries, pneumothorax) or those requiring immediate critical care admission.

### Data Extraction and Quality Assessment

Two reviewers (SL and MB) independently extracted data using Covidence. Extracted data included population demographics, intervention details, sample sizes, and outcome measures.

Methodological quality was assessed using the Mixed Methods Appraisal Tool (MMAT) (2018 version) [16], which is designed for reviews including diverse study designs. This review included many study designs, and as such did not allow use of other more widely used quality assessment tools such as the Cochrane Risk of Bias tool. Two reviewers (SL and MB) assessed each paper, with disagreements resolved by consensus. An overall quality score was not assigned, per MMAT guidance [17]; instead, a descriptive summary of quality is presented.

### Outcome Measures

The primary outcome was pain, as measured by the Visual Analogue Scale (VAS) or Numeric Pain Rating Scale (NPRS). Secondary outcomes were: rates of complications (e.g., pneumonia, delayed haemothorax), length of hospital stay (days), 30-day mortality, and additional opioid requirements (mg morphine equivalents).

### Data Analysis

Statistical analysis was performed using R software (v. 4.0.0) with the ‘metafor’ package. Data on participant numbers, mean and standard deviations were collected from papers where available for each of the study outcome measures. Where studies had reported median and Inter-quartile ranges, estimates of sample mean and standard deviation were derived from the sample size, median and interquartile ranges reported using the method outlined by Wan, et al. [7], to allow for further assessment to be completed across studies through meta-analysis. Meta-analysis was conducted where appropriate using a REML random effects model. Standardised Mean Difference (SMD) was calculated for continuous data (pain, length of stay, opioid requirement), and Risk Ratio (RR) was calculated for dichotomous data (complications), both with 95% confidence intervals (CIs).

Heterogeneity was assessed visually using forest plots and statistically using Higgins’ I^2^ and Chi^2^ (□^2^). Heterogeneity was classified as low (<40%), moderate (30-60%), substantial (50-90%), or considerable (75-100%) [18]. Sensitivity analyses were performed by removing studies with significant methodological differences to explore sources of high heterogeneity. The GRADE approach was used to assess the certainty of evidence for the primary outcome [19].

## Results

### Article Selection

Combining the initial and updated searches, 1015 studies were identified. After removing 451 duplicates, 564 titles and abstracts were screened. Of these, 47 papers were sought for full-text retrieval. One paper was not retrievable, and 46 were assessed for eligibility. Thirty-two papers were excluded (24 were editorial letters, single-case series or reviews, 7 used a different intervention, and 1 involved a different patient group).

Fourteen studies were included in the review. These comprised seven RCTs, one pilot study, two retrospective observational studies, three prospective observational studies, and one retrospective experimental trial. Nine studies contributed to the meta-analyses. The PRISMA flow diagram is shown in **Figure 1**.

**Figure 1.**
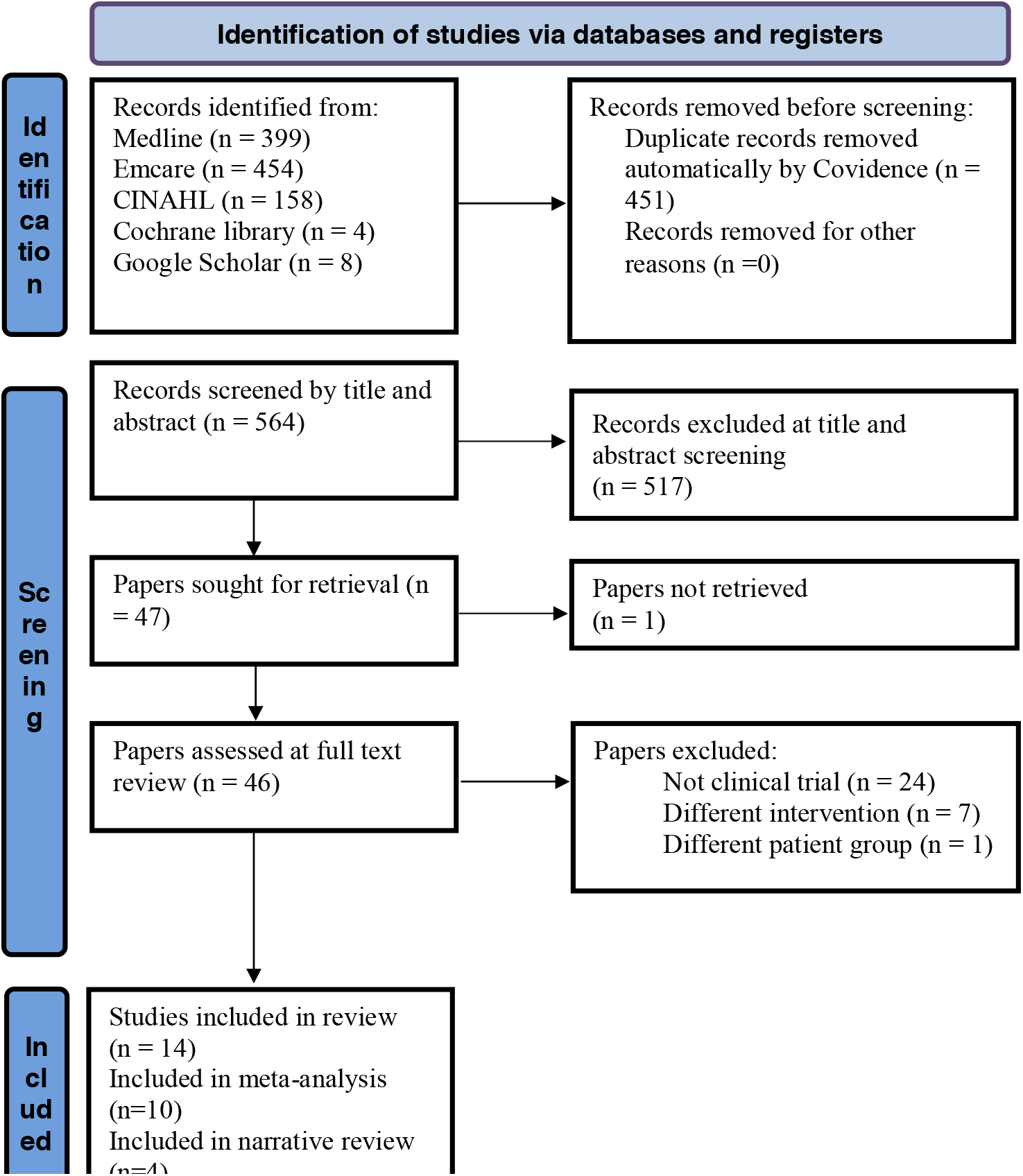
PRISMA flow diagram

### Methodological Quality and Demographics

The MMAT quality assessment is detailed in **Table 1**. All 14 included papers addressed a clear research question. Of the five RCTs, four used appropriate randomisation [5, 10, 20, 21]; one used a quasi-random allocation method based on odd/even participant number, which is considered at high risk of selection bias [11]. Assessors were not blinded in any RCT; however, this risk of bias is partly mitigated because the primary outcome (pain) was self-reported. Of the seven non-RCT studies, five did not adequately measure or adjust for key confounding variables [12, 13, 22-24].

**Table 1:** Quality Review of included studies using MMAT (2018)

| Criteria | Sareen<br>(2015)<br>[27] | Akca<br>(2019)<br>[11] | Cakmak<br>(2020)<br>[12] | Bakker<br>(2022)<br>[24] | Agir &<br>Cansun<br>(2025)<br>[14] | Srinivas<br>arangan<br>(2022)<br>[25] | Surdhar<br>(2023)<br>[23] | Serra<br>(2024)<br>[26] | Ramesh<br>(2024)<br>[21] | Partyka<br>(2024)<br>[5] | Sadauskas<br>(2024)<br>[22] | Perice<br>(2026)<br>[6] | Batome<br>n Kuimi<br>(2019)<br>[13] | Alar<br>(2021)<br>[20] |
| --- | --- | --- | --- | --- | --- | --- | --- | --- | --- | --- | --- | --- | --- | --- |
| S1. Clear research questions? | ✓ | ✓ | ✓ | ✓ | ✓ | ✓ | ✓ | ✓ | ✓ | ✓ | ✓ | ✓ | ✓ | ✓ |
| S2. Data allow to address question? | ✓ | ✓ | ✓ | ✓ | ✓ | ✓ | ✓ | ✓ | ✓ | ✓ | ✓ | ✓ | ✓ | ✓ |
| 2.1. Randomisation appropriate? |  | x |  | ✓ | ✓ |  |  |  | ✓ | ✓ |  | ✓ |  | ✓ |
| 2.2. Groups comparable at baseline? |  | ✓ |  | ✓ | ✓ |  |  |  | ✓ | ✓ |  | ✓ |  | ✓ |
| 2.3. Complete outcome data? |  | ✓ |  | x | ✓ |  |  |  | ✓ | x |  | ✓ |  | ✓ |
| 2.4. Outcome assessors blinded? |  | x |  | x | x |  |  |  | x | x |  | ✓ |  | x |
| 2.5. Participants adhered to intervention? |  | ✓ |  | x | ✓ |  |  |  | ✓ | x |  | ✓ |  | ✓ |
| 3.1. Participants representative? | ✓ |  | ✓ |  |  | ✓ | ✓ | ✓ |  |  | ✓ |  | ✓ |  |
| 3.2. Measurements appropriate? | ✓ |  | ✓ |  |  | ✓ | ✓ | ✓ |  |  | ✓ |  | ✓ |  |
| 3.3. Complete outcome data? | ✓ |  | ✓ |  |  | ✓ | x | x |  |  | ✓ |  | x |  |
| 3.4. Confounders accounted for? | x |  | x |  |  | ✓ | x | ✓ |  |  | x |  | x |  |
| 3.5. Intervention administered as intended? | ✓ |  | ✓ |  |  | ✓ | ✓ | ✓ |  |  | ✓ |  | x |  |

✓ = positive response; x = negative response; blank = not applicable

Participant demographics are summarised in **Table 2**. Across all 14 studies, the average participant age was 58.4 (±6.84) years, and 65.2% were male. The most common mechanisms of injury were road traffic accidents (35.5%) and falls from standing height (34.7%) [13, 20, 21, 25].

**Table 2:** Summary of demographics of included studies.

| Paper | Study type | Intervention | N (Interv) | N (Control) | Sex (M/F) (Interv) | Sex (M/F) (Control) | Age (Interv) | Age (Control) | Outcome Measure |
| --- | --- | --- | --- | --- | --- | --- | --- | --- | --- |
| Sareen (2015) [27] | Retrospective study | Kinesiotaping | 10 | 0 | 5 / 5 | / | 27-57 | / | NPRS |
| Akca (2019) [11] | Prospective randomised controlled study | Kinesiotaping | 16 | 14 | 12 / 4 | 9 / 5 | 50.1 | 49.4 | VAS |
| Cakmak (2020) [12] | Prospective Observational | Kinesioaping | 23 | 23 | 17 / 6 | 12 / 11 | 53.3 | 55.9 | VAS |
| Bakker (2022) [24] | Pilot randomised controlled trial | Kinesiotaping | 40 | 43 | 18 / 22 | 21 / 22 | 56.4 | 50.7 | NPRS |
| Agir & Cansun (2025) [14] | Randomised controlled trial | Kinesiotaping | 15 | 15 | 9/6 | 8/7 | 63.3 | 60.76 | VAS |
| Srinivasarangan (2022) [25] | Retrospective observational study | ESPB | 15 | 0 | 15 / 0 | / | 49.3 | / | NPRS + Peak Expiratory Flow Rate |
| Surdhar (2023) [23] | Prospective pilot Observational study | ESPB | 11 | 0 | UK | UK | 59.9 | / | NPRS |
| Serra (2024) [26] | Retrospective observational study | SAPB | 75 | 81 | 53 / 22 | 49 / 32 | 64.0 | 61.0 | NPRS + Opioid consumption + Number of adverse events |
| Ramesh (2024) [21] | Randomised controlled trial | ESPB | 23 | 23 | 21 / 2 | 19 / 4 | 49.5 | 51.5 | Amount of rescue analgesia + NPRS |
| Partyka (2024) [5] | Multicentre randomised controlled trial | SAPB | 103 | 104 | 67 / 36 | 62 / 42 | 71.0 | 71.0 | NPRS + Rate of pneumonia + Length of hospital stay + Opioid consumption |
| Sadauskas (2024) [22] | Prospective observational cohort study | SAPB | 15 | 23 | 10 / 5 | 10 / 13 | 58.0 | 64.0 | NPRS + expected change in IS volumes |
| Perice (2026) [6] | Prospective randomised controlled trial | SAPB | 19 | 19 | 12/7 | 13/6 | 67.9 | 69.8 | Pain, inspiratory capacity and cough (PIC) score |
| Batomen Kuimi (2019) [13] | Prospective Observational Cohort study | IS | 182 | 257 | 120 / 62 | 178 / 79 | 57.0 | 55.2 | Rate of delayed complications |
| Alar (2021) [20] | Prospective randomised controlled trial | IS | 57 | 57 | 40 / 17 | 37 / 20 | 58.4 | 54.2 | Length of hospital stay + Rates of complications |

*ESPB = Erector Spinae Plane Block; IS = Incentive Spirometry; SAPB = Serratus Anterior Plane Block; UK = Unknown*

### Primary Outcome: Pain

Twelve studies assessed pain: five kinesiotaping studies and seven chest wall regional anaesthesia studies [5, 11, 12, 21-27]. No incentive spirometry studies reported pain as an outcome.

#### For Kinesiotaping

five studies (n=182) were included in the meta-analysis for pain at days 3-4 post-intervention [11, 12, 24]. The pooled standardised mean difference favoured kinesiotaping over medication alone (SMD -1.61, 95% CI [-2.37 to -0.86]) (figure 2), although betweenLJstudy heterogeneity was considerable (I^2^ = 82.15%, p < 0.001). The study by Bakker et al. [24] included other injuries (e.g., clavicle fractures), differing from the other studies [11, 12, 33, 34]. When Bakker, et al. [24] was removed in the sensitivity analysis, the pooled effect remained in favour of kinesiotaping (SMD -1.87, 95% CI -2.65 to -1.08) and heterogeneity was reduced but remained substantial (I^2^= 73.22%, p=0.0074). The high heterogeneity may be due to possible variations in kinesiotaping methods; indeed, there is currently no consensus on the best application method.

**Figure 2.**
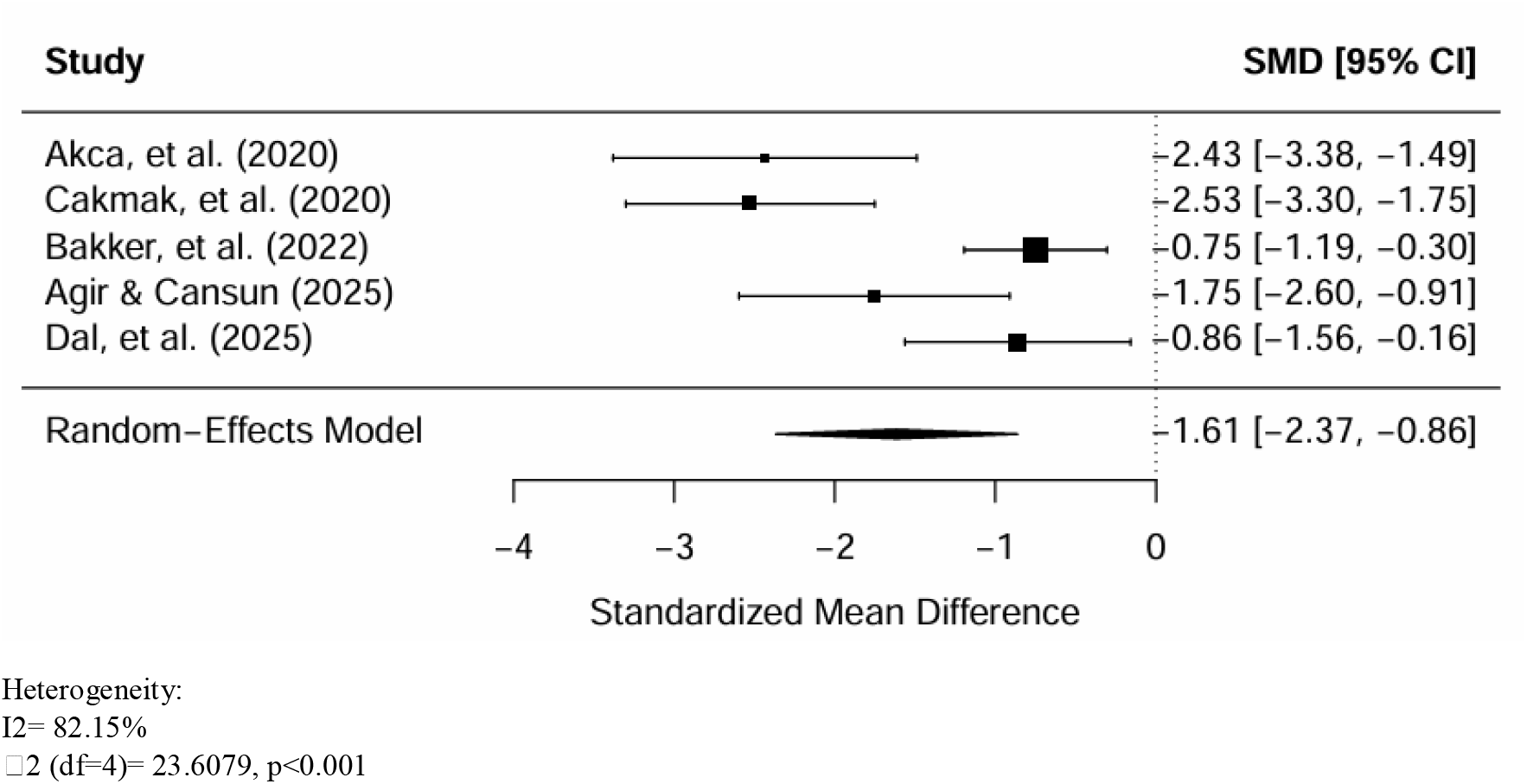
Forest plot presenting the effect of kinesiotaping + medication on pain compared to medication alone.

A fourth study (n=10), not included in the meta-analysis due to its design, found kinesiotaping immediately reduced pain on coughing (NPRS 7.2 to 4.7) and deep breathing (NPRS 5.7 to 4.4) [27]. Based on the GRADE approach, there was low-moderate certainty evidence for kinesiotaping reducing pain.

#### For single-shot chest wall regional anaesthesia (SAPB and ESPB)

Three studies (n=259) assessed pain at 120-180 minutes post-procedure and were included in the meta-analysis [21, 22, 26]. The pooled data demonstrated a significant reduction in pain favouring SAPB and ESPB over medication alone (SMD: -0.58, 95% CI [-1.15, -0.01]) (**Figure 3**). Heterogeneity was substantial (I^2^ = 71.94%, p = 0.046). The study by Sadauskas et al. [22] showed no effect, differing from the other two [5, 26].

**Figure 3.**
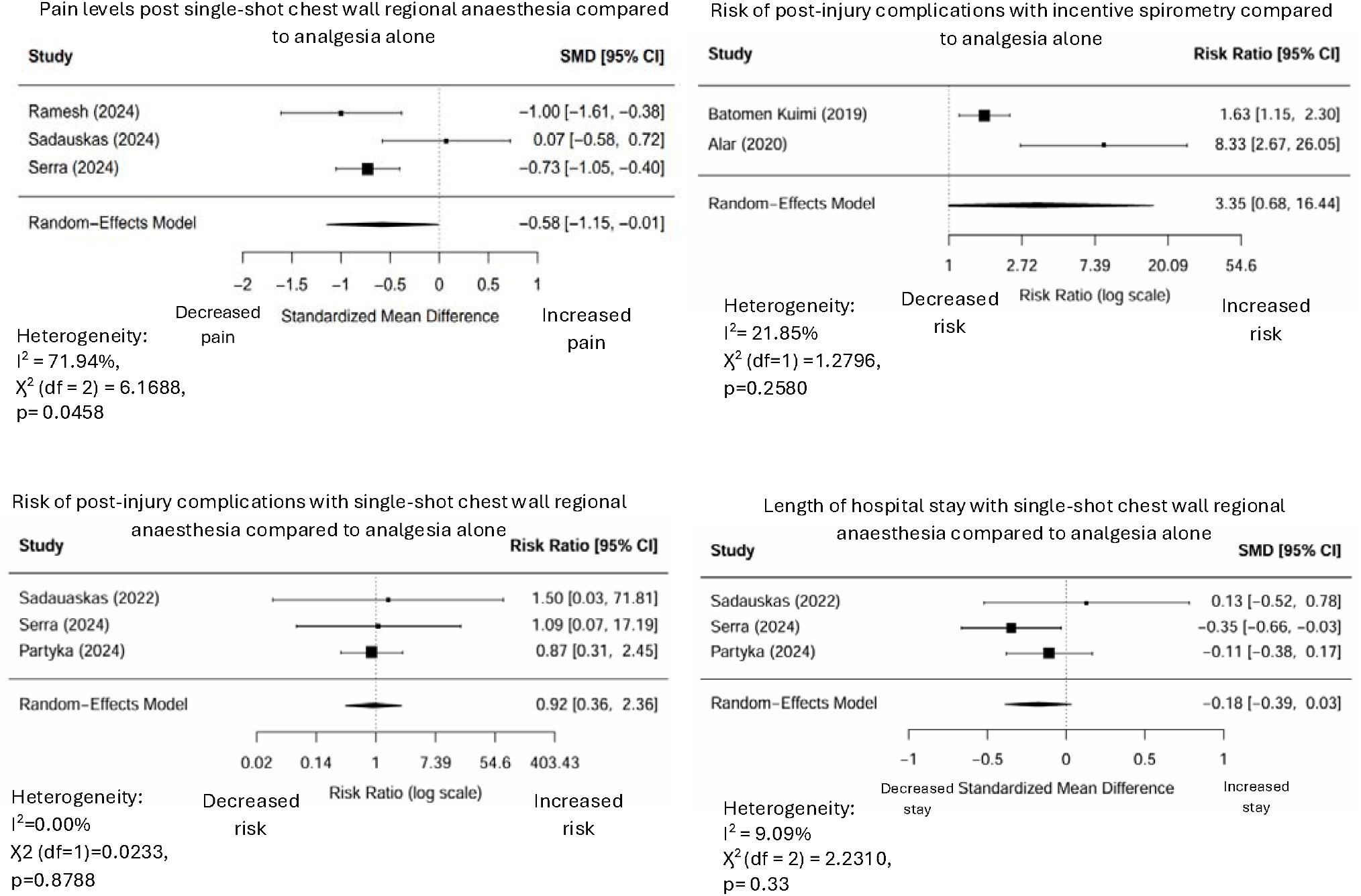
forest plots of meta-analysis outcomes

A sensitivity analysis removing Sadauskas et al. [22] resolved the heterogeneity (I^2^ = 0.00%) and strengthened the finding in favour of single-shot chest wall regional anaesthesia (SMD = -0.79, 95% CI [-1.07, -0.50]). The studies not in the meta-analysis also reported significant pain reduction post-block [5, 23, 25, 38]. Based on the GRADE approach, there was low-moderate certainty evidence for single-shot chest wall regional anaesthesia reducing pain.

### Secondary Outcomes

#### Risk of Complications

was assessed in incentive spirometry (2 studies) [13, 20] and single-shot chest wall regional anaesthesia (3 studies) [5, 22, 26] papers.

#### Incentive Spirometry

The pooled data (n=553) showed an increased risk of delayed haemothorax in the incentive spirometry group (RR: 3.35, 95% CI [0.68, 16.44]) (**Figure 3**). Heterogeneity was low (I^2^ = 21.85%). However, in both studies, the incentive spirometry groups had significantly more rib fractures at baseline (e.g., 3.0±2.0 vs 2.2±1.5 in Alar et al. [20]), a major confounding factor.

#### Single-shot chest wall regional anaesthesia

Three SAPB studies (n=416) assessed pneumonia rates. The pooled data showed no statistically significant difference between groups (RR: 0.92, 95% CI [0.36, 2.36]) (**Figure 3**). Heterogeneity was absent (I^2^ = 0.00%).

#### Length of Hospital Stay (LOS)

Three SAPB studies (n=416) assessed LOS [5, 22, 26]. The pooled data showed a small, non-significant reduction in LOS favouring the SAPB/ESPB group (SMD = -0.18, 95% CI [-0.39, 0.03]) (**Figure 5**). Heterogeneity was low (I^2^ = 9.09%).

#### Additional Opioid Requirement

Four single-shot chest wall regional anaesthsia studies (n=499) assessed opioid requirements in morphine milligram equivalents [5, 21, 22, 26]. Due to considerable heterogeneity among the included studies (I^2^= 97.39%, p<0.001), pooling the data for a meta-analysis was not clinically appropriate; therefore, these findings are presented as a narrative synthesis. Overall the evidence indicates a strong trend towards reduced opioid consumption in patients receiving regional anaesthesia. Three of the four studies demonstrated a distinct reduction in post-intervention opioid requirements for the intervention group compared to those receiving standard analgesia alone. The single study that did not report a benefit (Sadauskas, et al. [22]) was a significant outlier. In this cohort, the intervention group had a statistically significantly higher number of rib fractures at baseline compared to the control group, heavily confounding the results and likely masking any opioid-sparing effect of the regional block.

#### Mortality

No 30-day mortality events were reported in the included studies.

A summary of this review’s findings and certainty of evidence is shown below in **Table 3**.

**Table 3.** Summary of Findings Table.

| Certainty Assessment |  |  |  |  |  | Number of patients |  | Effect |  | Certainty |
| --- | --- | --- | --- | --- | --- | --- | --- | --- | --- | --- |
| Number of studies | Study Design | Risk of bias | Inconsistency | Indirectness | Imprecision | Intervention | Comparison | Relative (95% CI) | Absolute (95% CI) |  |
| Pain: Kinesiotaping (Follow-up: mean 4 days; assessed with VAS/NPRS; Scale from 0 to 10) |  |  |  |  |  |  |  |  |  |  |
| 5 | RCT | Not Serious | Serious [a] | Not Serious | Not Serious | 104 | 95 | - | **SMD 1.61 SD lower** (2.65 lower to 1.08 lower) | Low-moderate [a] |
| Pain: single shot chest wall regional anaesthesia (follow-up: range 120 mins to 180 mins; assessed with VAS/NPRS) |  |  |  |  |  |  |  |  |  |  |
| 7 | RCT | Not Serious | Serious [b] | Not Serious | Not Serious | 261 | 250 | - | **SMD 0.79 SD lower** (1.07 lower to 0.5 lower) | Low-moderate [b] |
| Risk of complications: Incentive Spirometry (assessed with relative risk) |  |  |  |  |  |  |  |  |  |  |
| 2 | RCT | Not Serious | Serious [c] | Not Serious | Very Serious [c] | 78/239 (32.6%) | 49/314 (15.6%) | **RR 3.35** (0.68 to 16.44) | **367 more per 1,000** (from 50 fewer to 1,000 more) | Low [c] |
| Risk of complications (pneumonia): single shot chest wall regional anaesthesia (assessed with relative risk) |  |  |  |  |  |  |  |  |  |  |
| 3 | RCT | Serious [d] | Not Serious | Not Serious | Not Serious | 7/152 (4.6%) | 8/168 (4.8%) | **RR 0.92** (0.36 to 2.36) | **4 fewer per 1,000** (from 30 fewer 65 more) | Low [d] |
| Length of hospital stay: single shot chest wall regional anaesthesia (assessed with relative risk) |  |  |  |  |  |  |  |  |  |  |
| 3 | Non-RCT | Not Serious | Not Serious | Not Serious | Not Serious | 261 | 250 | - | **SMD 0.18 SD lower** (0.39 lower to 0.03 higher) | Moderate [e] |
| Opioid Consumption: Single shot chest wall regional anaesthesia (assessed with Mg morphine) |  |  |  |  |  |  |  |  |  |  |
| 4 | Non-RCT | Serious [f] | Very Serious [f] | Not Serious | Not Serious | 267 | 231 | - | Narrative synthesis: 3 of 4 studies reported reduced consumption; data not pooled due to high heterogeneity | Low-moderate [f] |

CI: confidence interval; RR: risk ratio; SMD: standardised mean difference

## Discussion

This systematic review and meta-analysis suggest that for adult ED patients with rib fractures, kinesiotaping and single-shot chest wall regional anaesthesia (SAPB/ESPB) are potentially effective adjuncts for pain management, while the evidence for incentive spirometry reducing complications is poor and confounded.

### Principal Findings

The primary finding is that both kinesiotaping and single-shot chest wall regional anaesthesia provide a statistically and clinically significant reduction in pain compared to medication alone. Both interventions demonstrated large pooled effects on pain versus opioids alone (kinesiotaping SMD: -1.87; single-shot chest wall regional anaesthesia SMD: -0.79 post-sensitivity), where SMD >0.8 indicated substantial clinical benefit. These effects were robust after excluding heterogeneous outliers. This supports the use of both interventions as powerful, opioid-sparing analgesics in the ED.

Secondary findings were also promising for single-shot chest wall regional anaesthesia. We found a reduction in opioid consumption (SMD:-0.84 [-2.18, 0.50]) and a trend towards reduced length of hospital stay (SMD: -0.18 [-0.39, 0.03]). This suggests the superior pain control from single-shot chest wall regional anaesthesia may translate into tangible clinical benefits, supporting the “ED Alternatives to Opioids (ED-ALTO)” programme model [8].

Conversely, the findings for incentive spirometry were concerning. The meta-analysis indicated a potential increase in the risk of delayed haemothorax (RR 3.35[95%CI 0.68, 16.44). However, this result must be interpreted with extreme caution. In both included studies, the intervention groups had a significantly higher number of rib fractures at baseline [13, 20]. As fracture severity is a primary driver of complications [28], it is highly likely that this finding represents confounding by indication rather than a true harmful effect of the intervention. The data from these two studies are insufficient to draw any conclusion on the utility of incentive spirometry.

### Interpretation and Context

Heterogeneity in the primary analyses merits discussion. For kinesiotaping, Bakker et al. [24] attenuated the pooled effect due to the inclusion of concomitant shoulder and chest wall injuries. For single-shot chest wall regional anaesthesia, Sadauskas et al. [22] was the outlier (no benefit), likely due to a comparatively lower dosage of bupivacaine (0.25%) used for the SAPB, or baseline confounding, as the intervention group had more fractures. These factors highlight the need for standardised dosing and patient selection in future regional anaesthesia trials.

High heterogeneity in opioid consumption analysis (I^2^=97.39%) similarly precludes firm conclusions, though the effect was consistently in favour of single-shot chest wall regional anaesthesia in three of the four studies.

Our pain reduction findings for single-shot chest wall regional anaesthesia align with post-thoracic surgery evidence [10,29], but this is the first synthesis for an ED setting. Kinesiotaping results extend its established benefits in low back to chest wall injury [30], positioning it as a simple, non-pharmacological adjunct for use by ED clinicians and physiotherapists.

### Strengths and Limitations

This review’s strengths include a comprehensive search, adherence to PRISMA guidelines, prospective registration on PROSPERO registration, and dual-reviewer screening and quality assessment using the MMAT.

Limitations include the quality and quantity of available evidence, with only 12 studies meeting inclusion criteria (five RCTs). Many non-RCT’s failed to account for key confounders, limiting certainty, particularly for incentive spirometry. Significant heterogeneity was present in key analyses (pain and opioid use), though sensitivity analysis identified probable sources in study populations/protocols. The average participant age across studies was 57 years, so findings may not generalise to elderly patients (>65 years) who face the highest mortality risk from rib fractures [4] and opioid side effects

### Equity Considerations

1. **Country income level and health system context** Most included studies (n=9) were conducted in upper-middle or high-income countries with established emergency care systems, with only three studies originating from other settings. This may limit the applicability of findings to low-resource environments where access to imaging, ultrasound-guided regional anaesthesia, and postoperative care may differ substantially.
2. **Gender representation and sex-based differences** Eleven studies included both male and female participants, with one study including only male participants. Across all studies, males were consistently over-represented relative to females. None of the studies reported sex-stratified outcomes or explored sex-based differences in treatment response, despite evidence that females may experience greater post-fracture pain and higher complication rates (OR 5.6; 95% CI 1.1–29.6; p=0.043) [9]
3. **Socioeconomic factors and access to care** No study reported on participants’ socioeconomic status or other indicators of social disadvantage, and access to ultrasound-guided blocks or related resources was not examined. This absence of data limits understanding of how socioeconomic inequalities may influence access to, or benefit from, ultrasound-guided interventions for rib fractures.

Future research should adopt an intersectional approach, reporting race and ethnicity, sex, socioeconomic position, and care setting together, to better capture how overlapping social identities and structural factors influence access to rib fracture interventions and outcomes.

### Implications and Future Research

These findings support incorporating kinesiotaping and single-shot chest wall regional anaesthesia (SAPB/ESPB) into ED pathways for acute rib fracture management. Kinesiotaping represents a low-cost, low-risk intervention, while single-shot chest wall regional anaesthesia appears highly effective for patients with more severe pain or multiple fractures. Future research urgently requires high-quality RCTs to confirm these findings, particularly evaluating:

1. The true effect of incentive spirometry, using a protocol that controls for fracture severity.
2. The comparative effectiveness and duration of ESPB versus SAPB for this specific indication.
3. The effectiveness of these interventions as part of a wider bundle of care in the high-risk elderly population is being explored by the RELIEF trial for lidocaine patches [2].
4. The impact of these interventions on long-term outcomes, such as chronic pain and disability.
5. Inclusive recruitment across age, sex, ethnicity, and healthcare settings to support equitable strategies and address the current evidence gap [9]

## Conclusion

This systematic review and meta-analysis provide low-to-moderate certainty evidence that kinesiotaping and single-shot chest wall regional anaesthesia (SAPB and ESPB) significantly reduce pain in adult ED patients with rib fractures compared to standard opioid management. single-shot chest wall regional anaesthesia may also reduce total opioid consumption and length of hospital stay. Evidence for incentive spirometry is currently insufficient to support its use, as existing studies are confounded by fracture severity.

## Supporting information

Supplementary file 1 - search strategy

PRISMA Checklist

PROSPERO form

## Data Availability

All data produced in the present study are available upon reasonable request to the authors

## Declarations

### Availability of Data and Materials

The datasets generated and/or analysed during the current study are available from the corresponding author on reasonable request.

