## Supplementary file 1 - search strategy for "Kinesiotaping, single-shot chest wall regional anaesthesia, and Incentive Spirometry for Rib Fracture Management in the Emergency Department: A Systematic Review and Meta-Analysis"

Initial search 12th December 2024

Updated Search 8th June 2026

|  | Keyword | Medline | Emcare | CINAHL | Cochrane Library |
| --- | --- | --- | --- | --- | --- |
| 1. | Rib Fractures/ | 224 | 457 | 5 | 0 |
| 2. | (rib* and fracture*) | 818 | 588 | 12 | 0 |
| 3. | 1 or 2 | 818 | 588 | 12 | 0 |
| 4. | Chest wall injury* | 47 | 32 | 18 | 1 |
| 5. | 3 or 4 | 827 | 594 | 29 | 1 |
| 6. | Emergency Department* | 19,224 | 10,968 | 7,190 | 9 |
| 7. | “Accident and Emergency” | 168 | 3,263 | 25 | 0 |
| 8. | 6 or 7 | 19,342 | 11,016 | 7,202 | 9 |
| 9. | 5 and 8 | 48 | 47 | 2 | 1 |

|  | Keyword | Medline | Emcare | CINAHL | Cochrane Library |
| --- | --- | --- | --- | --- | --- |
| 1. | Rib Fractures/ | 3,910 | 1857 | 2,033 | 36 |
| 2. | (rib* and fracture*) | 8,963 | 6280 | 2.354 | 120 |
| 3. | 1 or 2 | 8,963 | 6280 | 2.354 | 120 |
| 4. | Chest wall injury* | 190 | 128 | 124 | 151 |
| 5. | 3 or 4 | 9,032 | 6315 | 2428 | 264 |
| 6. | Emergency Department* | 143,194 | 92,811 | 88,908 | 1842 |
| 7. | “Accident and Emergency” | 5,301 | 3206 | 2350 | 92 |
| 8. | 6 or 7 | 145,349 | 94,187 | 89,576 | 1842 |
| 9. | 5 and 8 | 351 | 407 | 156 | 3 |

‌
