## Supplementary material for "Kinesiotaping, single-shot chest wall regional anaesthesia, and Incentive Spirometry for Rib Fracture Management in the Emergency Department: A Systematic Review and Meta-Analysis": PROSPERO form

NIHR | National Institute for Health and Care Research

PROSPERO
International prospective register of systematic reviews

### The management of rib fractures following blunt chest wall trauma in adults presenting to the emergency department: a systematic review and meta-analysis

*Sophie Lenihan, Marc Barr, Nick Mani*

#### REVIEW TITLE AND BASIC DETAILS

##### Review title

The management of rib fractures following blunt chest wall trauma in adults presenting to the emergency department: a systematic review and meta-analysis

##### Review objectives

Does the use of taping, incentive spirometry or ultrasound guided nerve blocks reduce mortality rate, lower respiratory tract infection rate, length of hospital stay, and pain compared to standard care with oral medications alone in adult patients presenting to the emergency department with rib fractures following blunt chest wall injury.
Patients: Adults with rib fractures following blunt chest wall trauma
Intervention: ultrasound guided nerve blocks (single shot & catheter infusion, with or without perineural/IV dexamethasone) including serratus anterior plane (deep & superficial), erector spinae plane, paravertebral, taping, incentive spirometry
Comparison: Oral pain relief
Outcomes: mortality rate, lower respiratory tract infection rate, length of hospital stay, and pain

##### Keywords

blunt chest wall injury, emergency department, management, Rib fractures

#### SEARCHING AND SCREENING

##### Searches

Ovid MEDLINE, EMBASE, PROSPERO, Cochrane Library, CINAHL, and Scopus. A grey literature search will be included of Google Scholar, Embase, Emcare, BMJ Best Practice. References from selected studies will be identified during database search will be hand-searched to identify potential additional literature for consideration.
Studies with patients under 18 years, case reports, case series, editorials, letters, recommendations and

instructional articles, systematic reviews or meta-analyses are excluded.
No restriction to publication date, though only English language studies are considered.

##### Study design
Inclusion:
RCT
Observational (retrospective and prospective)
Cross-sectional studies of adult patients (18 years) with confirmed rib fractures presenting to emergency departments following blunt chest wall trauma
Exclusion:
International prospective register of systematic reviews
Studies that did not report the specific frequency of true positive, true negative, false positive and false
negative cases, or where this could not be calculated from other data provided, were excluded.

#### ELIGIBILITY CRITERIA

##### Condition or domain being studied
Rib fractures following blunt chest wall trauma in adults

##### Population
Adults presenting to the emergency department with chest injuries following blunt chest wall trauma.

##### Intervention(s) or exposure(s)
Ultrasound guided regional anaesthetic and/or kinesiology taping and/or incentive spirometry

##### Comparator(s) or control(s)
Standard care with analgesia (e.g. lidocaine topical patch, and oral or intravenous medications)

##### Context
Urgent/Acute/Emergency/Critical Care health care settings

#### OUTCOMES TO BE ANALYSED

##### Main outcomes
Mortality, lower respiratory tract infection, length of hospital stay or readmission, and pain

*Measures of effect*
Overall pooled effect

##### Additional outcomes
Standard error effect and confidence intervals

*Measures of effect*

#### DATA COLLECTION PROCESS

##### Data extraction (selection and coding)

Abstracts will be screened by two independent reviewers, and full text of the eligible studies will be reviewed
in the same manner based on the preformed inclusion and exclusion criteria. Where there was disagreement, a third independent review will be used to achieve the final consensus.
Characteristics of selected studies will be extracted using standardised pre-made table with the following
headings:
Author/
Publication Year/
Study Period
Setting/
Design/
Sample Size
Population
(Inclusion)
Exclusion
Intervention
(Index Test)
Target Condition
Definition
Comparison
(Reference Test)

##### Risk of bias (quality) assessment

The quality assessment of diagnostic accuracy studies (QUADAS-2) will be utilised to assess the quality of
the selected papers. This will involve following the guidance as set out on
https://www.bristol.ac.uk/population-health-sciences/projects/quadas/quadas-2/.
For each study there will be a completed bias form covering the domains of internal and external validity.
https://www.bristol.ac.uk/media-library/sites/quadas/migrated/documents/quadas2.pdf
Two independent reviewers will perform this task independently. Where there was disagreement, a third
independent review will be used to achieve the final consensus.

#### PLANNED DATA SYNTHESIS

##### Strategy for data synthesis

If appropriate meta-analysis (quantitative analysis) will be performed. This will be completed with statistical pooling of estimated overall mean effect of each treatment option: taping, incentive spirometry, and ultrasound guided regional anaesthetic. Standard error effects and confidence intervals will also be completed for this data. This will be performed using a random effects model.
The forest plots will be first visually examined for heterogeneity. Higgin’s I squared will be calculated to measure heterogeneity.
Statistical software 'R' will be utilised to calculate the above, and generate the tables.

In cases that quantitative analysis is not possible, synthesis of the findings will be provided with appropriate categorisation (qualitative analysis)
Two independent reviewers will perform this task independently. Where there was disagreement, a third independent review will be used to achieve the final consensus.

##### Analysis of subgroups or subsets

Standard error effect and confidence intervals

#### REVIEW AFFILIATION, FUNDING AND PEER REVIEW

##### Review team members


- Miss Sophie Lenihan, Chesterfield Royal Hospital NHS Foundation Trust
- Mr Marc Barr, Chesterfield Royal Hospital NHS Foundation Trust
- Dr Nick Mani, Chesterfield Royal Hospital NHS Foundation Trust


##### Review affiliation

Chesterfield Royal Hospital NHS Foundation Trust

University College London (UCL)

##### Funding source

Not applicable

##### Named contact

Sophie Lenihan. Chesterfield Royal Hospital, Chesterfield Road, Calow, S44 5BL


#### TIMELINE OF THE REVIEW

##### Review timeline

Start date: 01 November 2024. End date: 31 May 2025

##### Date of first submission to PROSPERO

17 October 2024

##### Date of registration in PROSPERO

27 November 2024

#### CURRENT REVIEW STAGE

##### Publication of review results

The intention is to publish the review once completed.The review will be published in English

##### Stage of the review at this submission

| Review stage | Started | Completed |
|---|---|---|
| Pilot work | | |
| Formal searching/study identification | | |

| Review stage | Started | Completed |
| --- | --- | --- |
| Screening search results against inclusion criteria | | |
| Data extraction or receipt of IP | | |
| Risk of bias/quality assessment | | |
| Data synthesis | | |

##### Review status

The review is currently planned or ongoing.

#### ADDITIONAL INFORMATION

##### PROSPERO version history

- Version 1.0 published on 27 Nov 2024

##### Review conflict of interest

None known

##### Country

England

##### Other registration details

##### Medical Subject Headings

Adult; Dexamethasone; Emergency Service, Hospital; Humans; Length of Stay; Motivation; Nerve Block; Pain; Rib Fractures; Spirometry; Thoracic Injuries; Thoracic Wall; Ultrasonography, Interventional

##### Details of any existing review of the same topic by the same authors

##### Disclaimer

The content of this record displays the information provided by the review team. PROSPERO does not peer review registration records or endorse their content.

PROSPERO accepts and posts the information provided in good faith; responsibility for record content rests with the review team. The owner of this record has affirmed that the information provided is truthful and that they understand that deliberate provision of inaccurate information may be construed as scientific misconduct.

PROSPERO does not accept any liability for the content provided in this record or for its use. Readers use the information provided in this record at their own risk.

Any enquiries about the record should be referred to the named review contact
